# Deep ensemble multitask classification of emergency medical call incidents combining multimodal data improves emergency medical dispatch

**DOI:** 10.1101/2020.06.26.20123216

**Authors:** Pablo Ferri, Carlos Sáez, Antonio Félix-De Castro, Javier Juan-Albarracín, Vicent Blanes-Selva, Purificación Sánchez-Cuesta, Juan M García-Gómez

**Affiliations:** Biomedical Data Science Laboratory (BDSLab), Instituto de Aplicaciones de las Tecnologías de la Información y de las Comunicaciones Avanzadas (ITACA), Universitat Politècnica de València (UPV), Valencia, Spain; Conselleria de Sanitat Universal i Salut Pública, Generalitat Valenciana (GVA), Valencia, Spain

**Keywords:** medical emergencies, emergency medical calls, emergency medical dispatch, deep learning, ensemble learning, multitask learning

## Abstract

The objective of this work was to develop a predictive model to aid non-clinical dispatchers to classify emergency medical call incidents by their life-threatening level (yes/no), admissible response delay (undelayable, minutes, hours, days) and emergency system jurisdiction (emergency system/primary care) in real time. We used a total of 1 244 624 independent incidents from the Valencian emergency medical dispatch service in Spain, compiled in retrospective from 2009 to 2012, including clinical features, demographics, circumstantial factors and free text dispatcher observations. Based on them, we designed and developed DeepEMC^2^, a deep ensemble multitask model integrating four subnetworks: three specialized to context, clinical and text data, respectively, and another to ensemble the former. The four subnetworks are composed in turn by multi-layer perceptron modules, bidirectional long short-term memory units and a bidirectional encoding representations from transformers module. DeepEMC^2^ showed a macro F1-score of 0.759 in life-threatening classification, 0.592 in admissible response delay and 0.757 in emergency system jurisdiction. These results show a substantial performance increase of 12.5%, 17.5% and 5.1%, respectively, with respect to the current in-house triage protocol of the Valencian emergency medical dispatch service. Besides, DeepEMC^2^ significantly outperformed a set of baseline machine learning models, including naive bayes, logistic regression, random forest and gradient boosting (α=0.05). Hence, DeepEMC^2^ is able to: 1) capture information present in emergency medical calls not considered by the existing triage protocol, and model complex data dependencies not feasible by the tested baseline models. Likewise, our results suggest that most of this unconsidered information is present in the free text dispatcher observations. To our knowledge, this study describes the first deep learning model undertaking emergency medical call incidents classification. Its adoption in medical dispatch centers would potentially improve emergency dispatch processes, resulting in a positive impact in patient wellbeing and health services sustainability.

## 1. Introduction

Emergency medical dispatch (EMD) involves the reception and management of requests for medical assistance in an emergency medical services system [1]. It comprises two main dimensions: call-taking, where emergency medical calls are received and incidents are classified according to their priority—triaged—and controlling, where the best available resources are dispatched to handle the event [2].

The call-taking process is generally managed by emergency medical dispatchers [3]. These mediators are in many cases non-clinical staff, trained with the essential knowledge of medical emergencies for the proper and efficient management of the incident [1,4]. Dispatchers usually follow a clinical protocol, established in the medical dispatch center, and periodically verified by medical supervisors [5].

However, despite preparation and the existence of triage protocols, assigning priorities to emergency medical call incidents (EMCI) is a challenging and stressful task for dispatchers, requiring constant concentration [6-8]. Additionally, there is always an inherent uncertainty on the real patient state, since the information of the event is gathered from telephonic interview processes. Furthermore, there are time constraints due to the incident priority or the need for tackling other incoming calls [9]. A wrong priority assignment derives either in insufficient medical attention or unnecessary resource deployment [10-12]. In consequence, EMCIs triage protocols are continuously revised and enhanced.

Many triage algorithms, such as the Emergency severity index [13], the Manchester triage system [14], the Canadian triage and acuity scale [15] or the Australasian triage scale [16], have been widely studied and enriched [17-20]. However, they are difficult to benchmark, deriving in no international agreement about their use for EMD [21]. Likewise, these algorithms depend on structured clinical information which is not always available during the call [22]. As such, improvements in EMD processes by redefining this sort of protocols are extremely costly and limited.

In the Valencian Community (Spain), the triage of EMCI is currently supported by an in-house triage protocol, based on a clinical decision tree, grounded on heavily structured clinical variables, e.g., *chest pain* (*yes* or *no*), collected throughout the interview in a sequential manner. Therefore, free text dispatcher observations, with higher expressiveness than structured data, cannot be automatically processed by the protocol, limiting its generalization to situations beyond the established guidelines.

The potential capability of deep learning to enhance EMCI classification through the provision of decision support to non-clinical dispatchers, was spotted by the Health Services Department of the Valencian region, aware of the potential of these models: deep learning is at the state of the art of machine learning in tasks involving complex types of data [23], e.g., high dimensional, unstructured, sequential, multimodal [24-27], such as those found in EMCI databases. Likewise, this and other machine learning tools have already been applied to tackle EMD challenges such as ambulance allocation [28-30], prediction of emergency calls volume [31], automatic stress detection of the caller [32], interpretable knowledge extraction [33], performance monitoring [34], cardiac arrest calls assistance [35] or triaging unconscious and fainting patients [36]. Therefore, we can argue that deep learning models are a feasible and promising technology to improve EMD through EMCI classification.

In this work, we develop and evaluate a deep learning model to provide decision support to non-clinical dispatchers in EMCI triage from the medical dispatch center of the Valencian region. Our model is designed to integrate the EMCI data collected during the call and carry out its classification. Despite of the existence of studies dealing with EMCI classification for specific disorders, as mentioned in the previous paragraph, to our knowledge, this is the first large-scale study undertaking a general EMCI classification trough deep learning.

## 2. Materials

### 2.1. Dataset

#### 2.1.1. Overview

A total of 1 244 624 independent EMCI of the Health Services Department of the Valencian Community, were compiled in retrospective from 2009 to 2012. The Health Services Department board of the Valencian Community approved the data use for this project, removing before their analysis any information that may disclose the identity of the person.

These EMCI data included during-call and after-call data. We categorized the data variables as structured—fixed fields—and unstructured—open fields—as well as stationary—with no implicit order—and sequential—with an implicit order (Figure 1).

**Figure 1.**
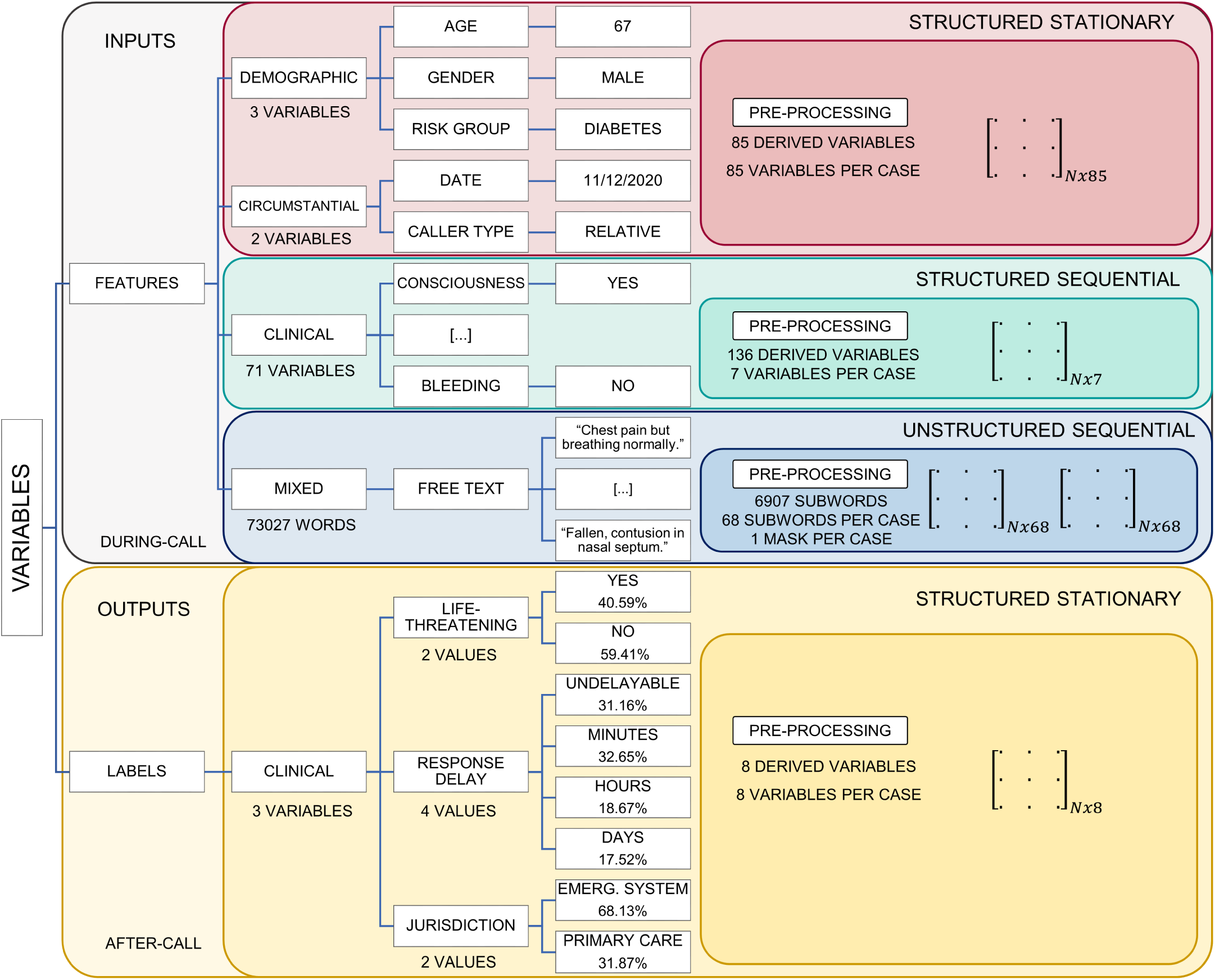
Dataset variables arranged by type. Names and cardinality, before and after pre-processing (derived variables), are presented, indicating how many variables—or subwords, when referring to text features—are available per case after pre-processing. Examples for their values are also included. Class frequencies for each output label are also reported. N is equal to the 722 270 EMCI used in the study.

#### 2.1.2. During-call data

During-call data (Figure 1 top) are recorded during the emergency medical call. These data consist of demographics, circumstantial factors, clinical features— collected throughout the triage tree navigation—and free text dispatcher observations:

Demographics data—structured and stationary— include age, gender and risk group variables. Age is a numerical discrete feature, gender is a categorical binary variable (*male, female*) and risk group is a categorical multiclass variable—with multiple possible values, such as *asthmatic, allergic, cardiac, diabetic, neoplastic*, etc.

Circumstantial factors data—structured and stationary—include date and caller type variables. The latter consists on a categorical multiclass variable, keeping information about the person or institution which made the emergency medical call and taking values such as *police, red cross, the patient, a relative*, etc.

Clinical variables data—structured and sequential— include features providing relevant medical information. They are collected in a sequential manner during the call, registering a subset of them, from the total 71 variables available. A full list including all these variables is available in Table 1. These variables are categorical, presenting one possible value or multiple ones. An example of how four clinical variables and their values are registered during an emergency medical call could be: *previous trauma, yes*; *hemorrhage, yes*; *bleeding site, rectal bleeding*; *consequences of the clinic, severe blood loss*.

**Table 1.**
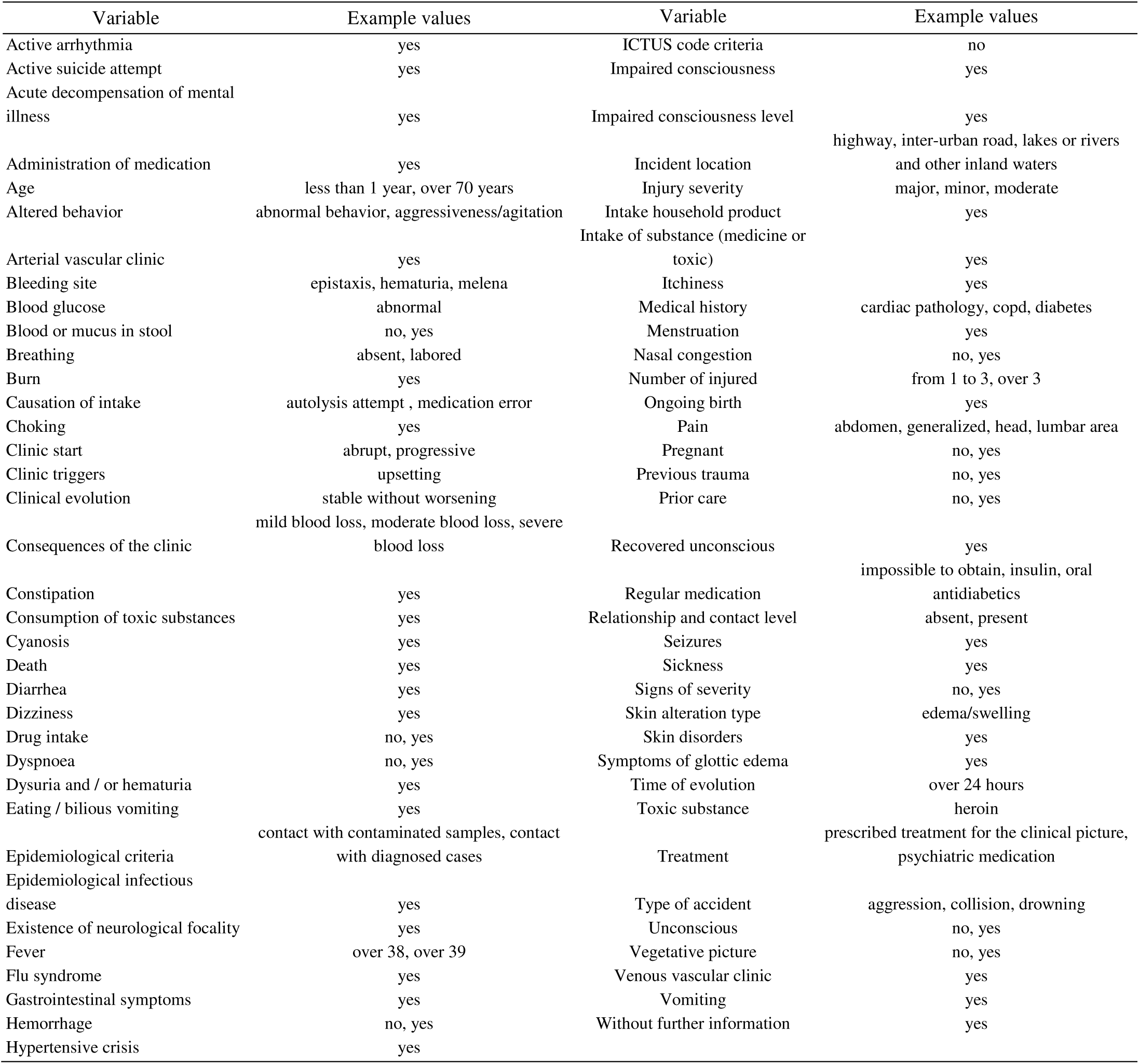
Clinical variables with some of their example values. Certain variables have just one possible associated value, while others may exhibit multiple values. To ease presentation, example values are limited to three in this table.

Finally, free text dispatcher observations— unstructured and sequential—consist on short sentences, written during the call and providing additional relevant information which cannot be recorded in a structured manner. The language in which they are written is Spanish. Examples of two free text dispatcher observations bound each one to a different event are (translated into English): *according to the caller epileptic crisis, he has drunk and taken pills, he is half-conscious with half-closed eyes*; *patient bleeds abundantly from the head after falling at home, they have just found it in a pool of blood*.

#### 2.1.3. After-call data

After-call data are recorded at a time after the call and used to derive EMCI classification labels, since they provide reliable up-to-date information about the real patient state. These data include: posterior physician diagnosis, standardized by International classification of diseases codes [37], such as *syncope* (ICD 780.2) or *acute myocardial infarction* (ICD 410); maneuvers and procedures indicating if the patient was *intubated, reanimated, sedated, received surgery*, etc.; and hospitalizations and urgency stays with information about the department where the patient was treated, the amount of time he stayed there and his discharge code.

#### 2.1.4. Labels derivation

We transcribed the information contained in after-call data to three different and complementary EMCI classification labels (Figure 1 bottom): life-threatening level (yes/no), admissible response delay (undelayable, minutes, hours, days) and emergency system jurisdiction (emergency system/primary care). The mapping between after-call data and EMCI classification labels was established by a panel of 17 physicians from the Health Services Department of the Valencian Community, using a Delphi methodology [38].

#### 2.1.5. Data quality assessment and inclusion criteria

To ensure the highest reliability of the model training data, we performed and reported a data quality analysis on the included data [39]. The analysis included the assessment of data quality dimensions of completeness and consistency, as well as temporal and multi-source variability [40-42]— changes in the statistical distributions of data over time or among sources, respectively. The main findings included: approximately 30% of data with at least one missing label; and outlying distributions in some dispatchers, especially those with less than 100 calls.

According to these results, we considered, for the next stages of our work, those EMCI which after-call data were fully available, and which during-call data were registered by non-novice dispatchers—dispatchers with more than 100 calls managed. The final working dataset size comprised 722 270 EMCI.

### 2.2. Framework

The implementation language was Python 3.7.3 [43], making use of libraries Pandas [44], NumPy [45], and Fuzzywuzzy [46], for data pre-processing and Sklearn [47], Pytorch (version 1.4.0) [48], Hugginface transformers [49] and Hyperopt [50] for modeling.

## 3. Methods

### 3.1. Data pre-processing

Depending on variable type, different pre-processing techniques were applied, mapping the original data to a matrix representation to be used for the deep learning model (Figure 1 right, highlighted pre-processing blocks):

Age, a structured stationary discrete ordinal variable, was mapped to a fuzzy [51] representation trough piecewise linear functions [52]. These membership functions, represented in Figure 2, were validated by physicians of the Health Services Department of the Valencian Community. This smoothing transformation was carried out to avoid sharp transitions derived from grouping in a small set of categories discrete ordinal variables with high cardinality in their values.

**Figure 2.**
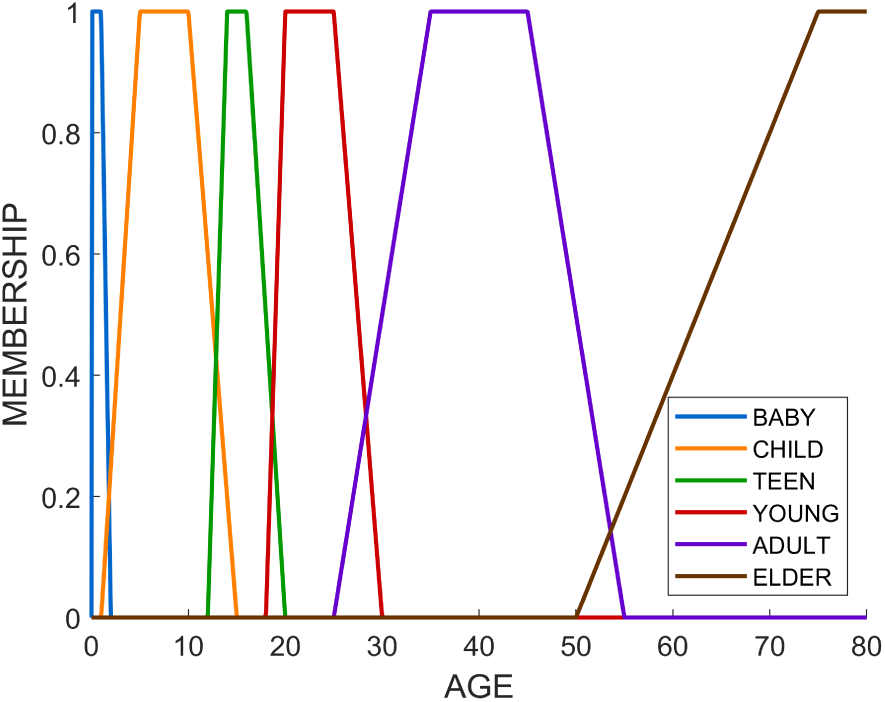
Piecewise linear functions representing age group membership

Gender, risk group and caller type, structured stationary categorical variables, were one-hot encoded while several variables were derived from the date variable: weekday, month, if the day was or not a weekend day and if the day was or not was a bank holiday. These resulting variables, also structured stationary categorical variables, were one-hot encoded too.

Regarding the clinical variables, structured sequential variables, each variable-value pair was converted to an integer, conforming then, sequences of integers that were pre-padded afterwards, to ensure sequences of fixed length [53]. This length was equal to 7, since in more than 99% of the incidents reported, the number of clinical variables collected was equal or lower than 7.

Spelling correction processes by means of fuzzy string matching [54] were applied to the free text dispatcher observations, unstructured sequential variables, to reduce vocabulary dimensionality and noise. Besides, subword tokenization with WordPiece was carried out to reduce vocabulary size [55]. To ensure sequences of fixed length while keeping information about the original sequences lengths, post-padding and attention mask generation were conducted. The padding length was set in 68, since in more than 99% of the incidents reported, the number of subwords written was equal or lower than 68.

Finally, labels, structured stationary categorical data, were one-hot encoded, deriving in a label matrix of 8 columns, each one associated with a specific label-class pair.

### 3.2. Data splitting and sampling

To evaluate model performance and tune hyperparameters without any bias, data were iteratively and randomly split into six subsets (Figure 3) [56]. First, data were randomly split into two disjoint *design* and *test sets*, with 80% and 20% proportions respectively. Next, the *design* set was randomly divided again into a *training* and a *validation set*, with 80% and 20% proportions. Finally, a sampling step was performed taking 100000 elements to define a *training* and a *validation sample*.

**Figure 3.**
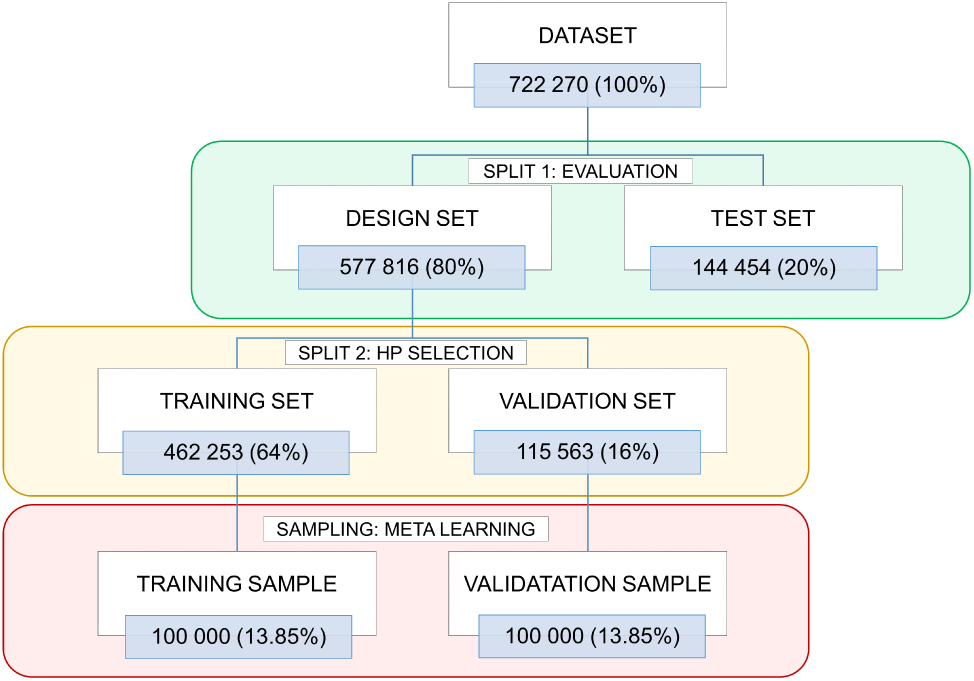
Data splitting and sampling. The number of data of each partition, along with its percentage respect the total number of data, are provided. Abbreviations: HP, hyperparameter.

### 3.3. Deep neural network design

The problem of classifying EMCI combining multimodal data was divided into four subproblems: three EMCI classification problems taking as inputs for each one EMCI data from the same type—structured stationary, structured sequential and unstructured sequential—and a last EMCI classification problem taking as inputs inner outputs obtained from the solution of the prior problems. To solve these four challenges, four deep learning (DL) subnetworks were developed: the *Context subnetwork* (ConNet), the *Clinical subnetwork* (CliNet), the *Text subnetwork* (TextNet) and the *Ensemble subnetwork* (EnsNet). Finally, once trained, they were combined in a single global modular neural network model [57].

Likewise, as the life-threatening, response delay and jurisdiction labels provide different but related information, e.g., a life-threatening situation implies a low admissible response delay, a multitask learning [58] paradigm was followed, to exploit these label dependences. To promote training efficiency and regularization while reducing the number of subnetworks parameters, a hard parameter sharing approach [59] was adopted. Hence, each of the four developed subnetworks presented a task-shared block—same set of parameters for all label prediction tasks—and a task-specific block—specific set of parameters for each label prediction task.

The ensemble of the four multitask subnetworks defined DeepEMC^2^—**Deep E**nsemble **M**ultitask **C**lassifier for **E**mergency **M**edical **C**alls—the global and definitive DL model.

Next, we describe in detail each of the subnetworks integrated in DeepEMC^2^, supported by Figure 4:

**Figure 4.**
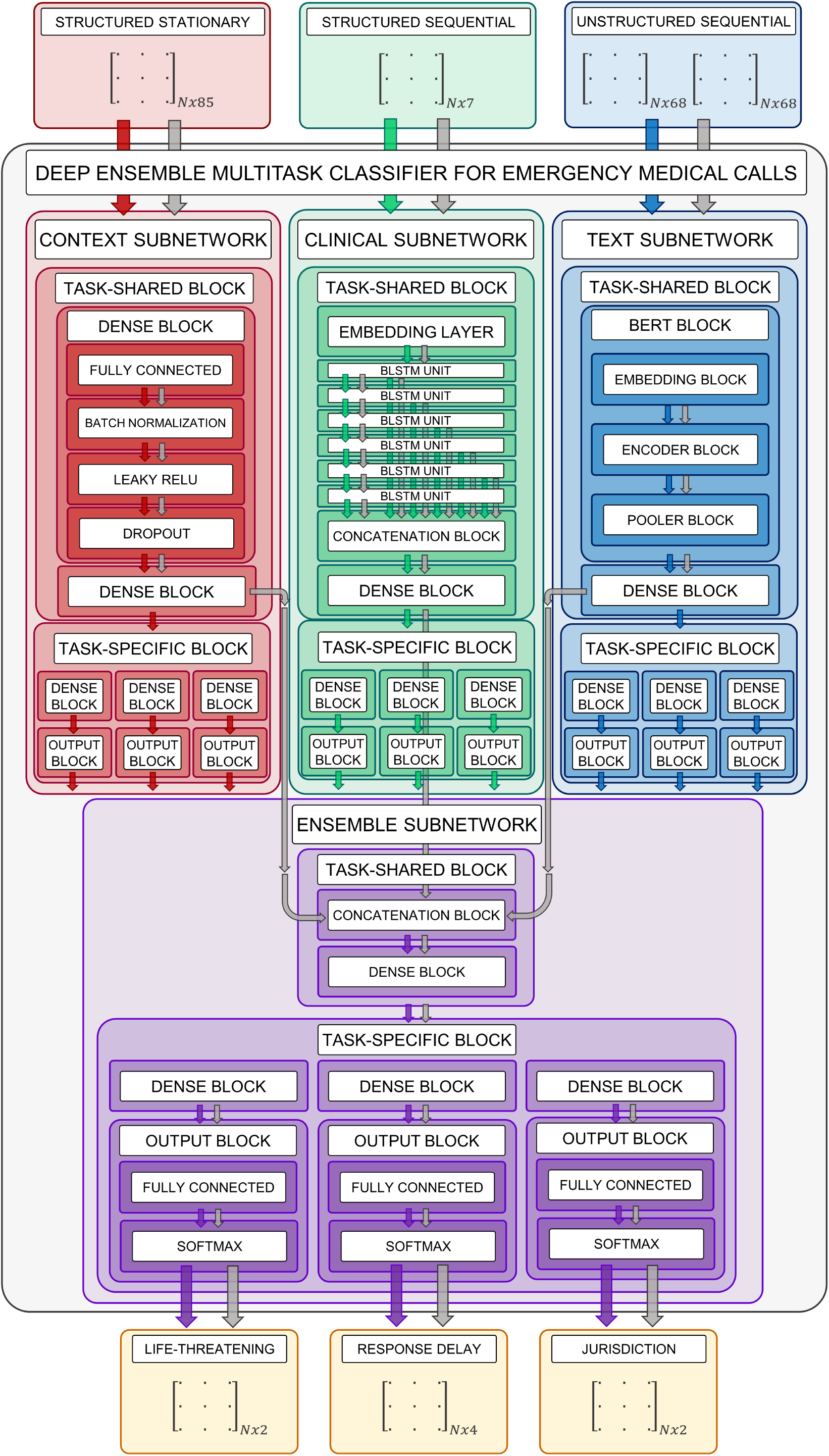
DeepEMC^2^—**Deep E**nsemble **M**ultitask **C**lassifier for **E**mergency **M**edical **C**alls—architecture, including its constituting subnetworks—the *Context subnetwork*, the *Clinical subnetwork*, the *Text subnetwork* and the *Ensemble subnetwork*. Arrows indicate the forward propagation direction, for each subnetwork, as well as the global network (DeepEMC^2^), colored according to the particular neural network they refer.

The *Context subnetwork* (Figure 4 left) deals with the demographics and circumstantial factors bound to an EMCI. It consists on a multi-layer perceptron (MLP) [60] due to its adequateness to model structured and stationary data, composed by dense and output blocks. A dense block integrates a fully connected layer [61] a batch normalization layer [62] to manage internal covariate shift, a leaky ReLU [63] activation function to avoid vanishing and exploding gradients, while preventing dead neurons issues [64] and a dropout layer [65] to prevent neuron co-adaptation. An output block is composed by a fully connected layer and a softmax activation function, to dispose of a normalization score— between 0 and 1—for each class of each predicted label.

The *Clinical subnetwork* (Figure 4 center) deals with the clinical features collected during the call. It consists on a recurrent model, since clinical features are notified in a sequential manner, being their recording order potentially informative. It is composed by an embedding layer [66], which compresses the sparse input space into a smaller and dense one; a stack of multiple bidirectional long short-term memory (BLSTM) [67] units, which capture long-term dependences far better than standard recurrent models; multiple skip connections [68] across the BLSTM units, to reduce the risk of losing relevant information during BLSTM propagation; a concatenation block—concatenates the outputs of these skip connections—and a MLP module, integrated by dense and output blocks, to act as an intermediary between the multiple BLSTM outputs and the final label predictions.

The *Text subnetwork* (Figure 4 right) deals with the free text dispatcher observations—unstructured and sequential—written during an EMCI. It is composed by a bidirectional encoding representations from transformers (BERT) [69] block, since this model is at the state of the art in natural language processing tasks, including text classification, and a MLP module, to relate BERT outputs with label outputs. The BERT clock is comprised in turn by an embedding block, an encoder block [70], and a pooler block, while the MLP component is constituted by dense and output blocks.

The *Ensemble subnetwork* (Figure 4 bottom) integrates inner outputs from the ConNet, the CliNet and the TextNet to generate the final outputs of DeepEMC^2^. It consists of a concatenation block with a MLP component, composed by dense and output blocks. The inputs of the concatenation block are the outputs of the last layer of the dense block prior to the task-specific block of each one of the former subnetworks. It takes these inner outputs, and not the final output scores since these last values aggregate tons of information in just a small set of scalar values; hence, the modeling potential of the inner outputs is higher.

### 3.4. Parameter tuning

Subnetworks were trained in a constructive modularized manner [57], so they were independently trained and assembled later as loosely coupled models. The optimizer selected for that was ADAM [71], given its learning adaptability, noisy gradients management and learning process stability [72,73]. A term of weight decay [74] was included in the parameters upgrading rule expression, to promote regularization. Likewise, it was followed a mini-batch upgrading approach [75], computing gradients with backpropagation [76] and backpropagation through time [77]. The objective function was a cross-entropy [78] loss (CEL). For each subnetwork, three CEL were calculated—one per label—averaged afterwards and finally backpropagated to carry out the parameter tuning process. Layers with leaky ReLU activation functions were initialized with Kaiming initialization [79], while softmax activation function layers were initialized with Xavier’s initialization [80].

### 3.5. Hyperparameter tuning

The influence of hyperparameters over subnetworks performance was carefully considered in this work, in order to maximize the attainable outcomes. The hyperparameters studied were related with subnetworks architecture and optimizer settings. These hyperparameters, as well as its definitive (*optimal*) values are presented in Table 2.

**Table 2.** Subnetworks hyperparameters with their definitive values obtained after carrying out the multi-step hyperparameter tuning process.

| Hyperparameters | Deep learning model subnetworks |  |  |  |
| --- | --- | --- | --- | --- |
|  | Context subnet | Clinical subnet | Text subnet | Ensemble subnet |
| Embedding dimension | - | 8 | 96 | - |
| DB hidden layers | 1 | 1 | - | 1 |
| DB neurons per layer | 64 | 64 | - | 256 |
| DB dropout | 0.1 | 0.1 | - | 0.25 |
| BLSTM layers | - | 6 | - | - |
| BLSTM neurons | - | 64 | - | - |
| BLSTM dropout | - | 0 | - | - |
| Encoder attention heads | - | - | 3 | - |
| Encoder layers | - | - | 3 | - |
| Encoder neurons per layer | - | - | 96 | - |
| Encoder dropout | - | - | 0.1 | - |
| TSB layers | 2 | 2 | 2 | 2 |
| TSB neurons | 64 | 64 | 64 | 64 |
| Batch size | 128 | 64 | 64 | 128 |
| Learning rate | 0.0005 | 0.0005 | 0.0005 | 0.0001 |
| Weigth decay | 0.00005 | 0.00005 | 0.0005 | 0.0005 |

Abbreviations: DB, dense blocks; TSB, task-specific blocks.

Hyperparameters were tuned following a multi-step strategy (Figure 5):

**Figure 5.**
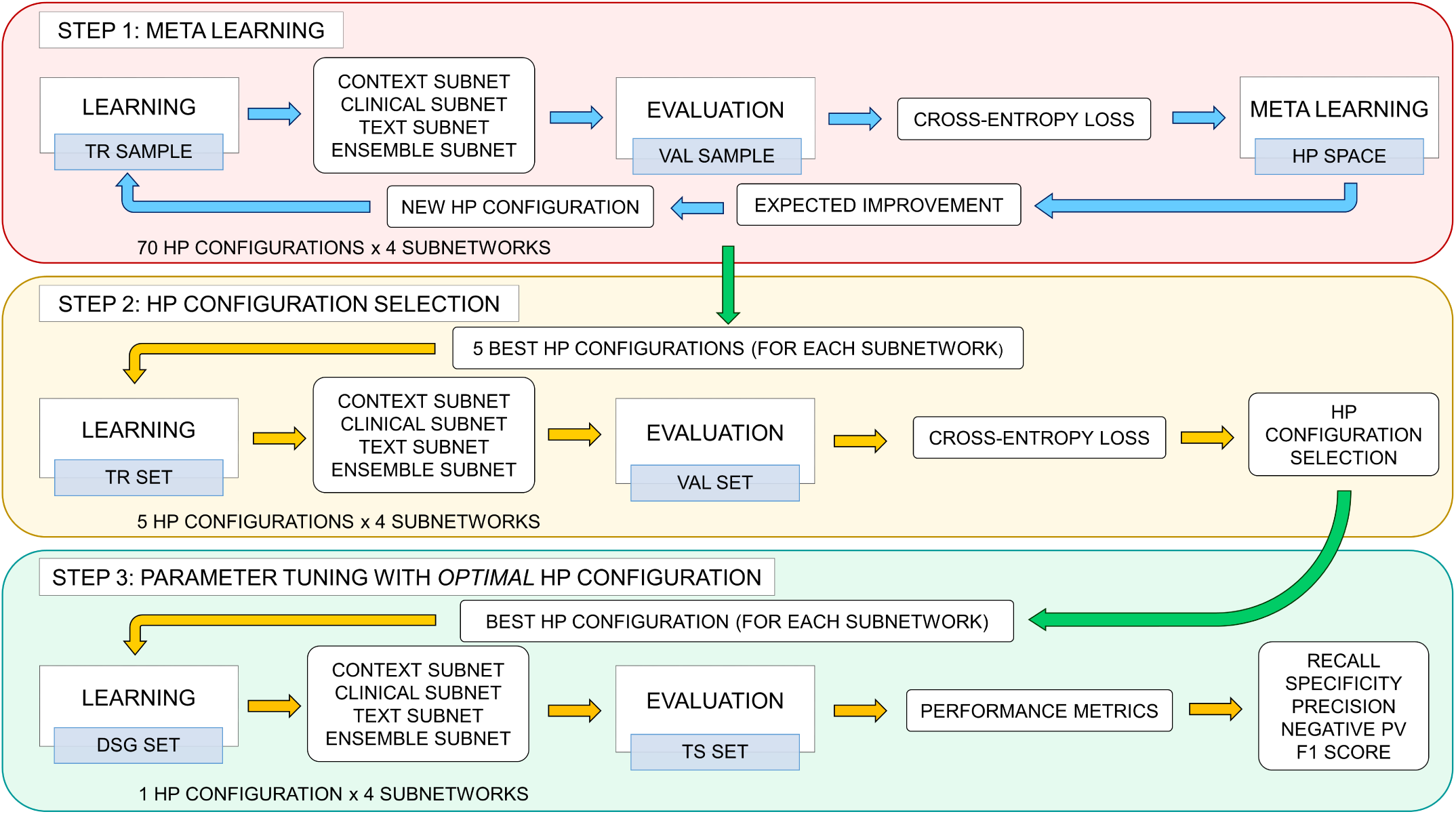
Multi-step hyperparameter tuning strategy. Yellow arrows imply unidirectionality, while blue arrows stand for a feedback loop, both inside a hyperparameter optimization step. Green arrows denote unidirectionality across hyperparameter optimization steps. Abbreviations: HP, hyperparameter; TR, training; VAL, validation; DSG, design TS, test.

The first step involved an automatic active learning [81] hyperparameter optimization process (Figure 5 top): four surrogate models—one per subnetwork—based on tree-structured parzen estimators [82], learned the conditional probability distribution of subnetworks hyperparameters given their associated CEL. Aiming to maximize the Expected Improvement [83] of the CEL, new hyperparamter configurations were iteratively sampled from the surrogate models, being upgraded after each training loop. Thereby, 280 different subnetworks—70 hyperparameter configurations times four subnetworks—were trained and evaluated in the *training* and *validation samples*, respectively.

Next, the best hyperparameter configurations proposed by the surrogate models were selected (Figure 5 middle). To prevent overfitting, the best five hyperparameter configurations for each subnetwork were taken to retrain and validate the subnetworks, in the *training* and the *validation set*, respectively, obtaining a total of 20 models trained in this step. Then, the CEL was obtained for each of them and those hyperparameter configurations with the best value—lowest validation CEL—were considered as the *optimal* hyperparameter configuration.

Finally, the *optimal* hyperparameters were used to retrain the four subnetworks using the whole *design set*, to ensure a proper exploitation of the data (Figure 5 bottom). Once trained, its integration into a single architecture defined DeepEMC^2^—the global network—evaluated later in the *test set*.

### 3.6. Evaluation

#### 3.6.2. In-house triage protocol and baseline models

First, to assess if DeepEMC^2^ provides an improvement in EMCI classification respect the existing clinical rules, performance metrics were obtained for the current in-house triage protocol of the Valencian emergency dispatch service.

Second, to compare the performance of the DL model respect well-known machine learning models in EMCI classification, we trained and evaluated the following baseline models:

- Multinomial naive bayes (NB) [84]: including a term of additive Laplace smoothing [85].
- Logistic regression (LR) [86]: including a penalty term for L2 regularization [87] and resorting to L-BFGS [88] as optimizer algorithm.
- Random forest (RF) [89]: considering Gini impurity as splitting criterion [90], while assembling a total of 300 tree estimators whose maximum depth was equal to 50, being these *optimal* values determined via hyperparameter tuning procedures.
- Gradient boosting (GB) [91]: considering mean squared error with improvement score by Friedman [91] as splitting criterion, with a total of 300 tree estimators whose maximum depth was set in 5, being these *optimal* values determined by hyperparameter tuning processes.

Notably, the input data for these baseline models had to be adapted to be processed by them. Clinical variables were one-hot encoded instead of being fed as sequences of integers. Regarding free text observations, once spelling correction processes, subword tokenization and sentence truncation were carried out, subwords were one-hot encoded.

#### 3.6.3. Metrics

Performance metrics were obtained in the *test set* (144 454 independent EMCI) for each label prediction task and each model trained—we recall here that EnsNet outputs are the same as DeepEMC^2^. The evaluation metrics included accuracy, recall, precision and F1-score [92,93]. For binary labels (life-threatening, jurisdiction), recall, precision and F1-score were referencing the interest class—life-thread and emergency system jurisdiction. Regarding the multiclass label (response delay), recall and precision were calculated for each class and then averaged following a macro approach. Likewise, for all labels, macro F1-score [92,93] was computed, to dispose of a balanced multiclass performance descriptor—not influenced by class frequencies. Finally, for all metrics, 95% confidence intervals were calculated by 1000 bootstrap samples [94] extracted from the test set.

Metrics were calculated in the *test set*, for the protocol, the baseline models—naive bayes, logistic regression, random forest and gradient boosting—and the DL models developed—the ConNet, the CliNet, the TextNet and DeepEMC^2^. We recall here that, although DeepEMC^2^ is the definitive DL model which takes into account input data globally, results referring its constituting subnetworks, contrasted with baseline models trained with the same type of input data of each subnetwork, are also reported, to analyze the contribution of each set of inputs to the global model and where DL provides a substantial gaining over the other kind of models.

Likewise, percentage differences between DeepECM^2^ and the protocol are also reported, as well as percentage differences between DeepECM^2^ and the best baseline model—that baseline model with the best balanced multiclass performance—which has been measured in our work in terms of macro F1-score.

## 4. Results

Tables 3, 4 and 5 show the classification performance results for the life-threatening level, admissible response delay and emergency system jurisdiction labels, respectively.

**Table 3.** Performances of the in-house triage protocol, baseline models and deep learning models in life-threatening prediction (test set). Bootstrapped 95% confidence intervals are shown between brackets. Percentage differences between DeepEMC^2^—the global deep learning model—and the protocol ΔP (%), along with percentage differences between DeepEMC^2^ and the best baseline model ΔBM (%)—highest F1-score and F1-score^MACRO^—are also reported.

| Model | Life-threatening level (yes/no) |  |  |  |  |
| --- | --- | --- | --- | --- | --- |
|  | Single-class metrics (yes) |  |  | Two-class metrics (yes/no) |  |
|  | Recall | Precision | F1-score | Accuracy | F1-score<sup>MACRO</sup> |
| Protocol | 0.644 [0.641, 0.647] | 0.547 [0.544, 0.551] | 0.592 [0.589, 0.595] | 0.639 [0.637, 0.641] | 0.634 [0.632, 0.636] |
| Context NB | 0.407 [0.404, 0.410] | 0.563 [0.559, 0.567] | 0.472 [0.469, 0.475] | 0.631 [0.629, 0.633] | 0.594 [0.592, 0.596] |
| Context LR | 0.411 [0.407, 0.414] | 0.577 [0.573, 0.581] | 0.480 [0.476, 0.483] | 0.638 [0.636, 0.640] | 0.601 [0.599, 0.604] |
| Context RF | **0.465 [0.462, 0.469]** | 0.526 [0.522, 0.529] | 0.494 [0.491, 0.497] | 0.612 [0.610, 0.614] | 0.590 [0.588, 0.592] |
| Context GB | 0.428 [0.425, 0.432] | **0.588 [0.584, 0.592]** | 0.495 [0.492, 0.499] | **0.646 [0.644, 0.648]** | 0.611 [0.609, 0.613] |
| Context DL | 0.440 [0.436, 0.443] | 0.583 [0.579, 0.587] | **0.501 [0.498, 0.504]** | 0.644 [0.642, 0.647] | **0.613 [0.610, 0.615]** |
| Clinical NB | 0.732 [0.729, 0.735] | 0.550 [0.547, 0.553] | 0.628 [0.625, 0.630] | 0.647 [0.645, 0.650] | 0.646 [0.644, 0.649] |
| Clinical LR | 0.752 [0.750, 0.755] | **0.586 [0.583, 0.589]** | 0.659 [0.656, 0.661] | 0.683 [0.681, 0.685] | 0.682 [0.680, 0.684] |
| Clinical RF | 0.764 [0.761, 0.767] | 0.585 [0.583, 0.589] | 0.663 [0.661, 0.665] | **0.684 [0.682, 0.686]** | **0.683 [0.681, 0.685]** |
| Clinical GB | 0.763 [0.760, 0.766] | 0.585 [0.583, 0.589] | 0.663 [0.660, 0.665] | **0.684 [0.682, 0.686]** | **0.683 [0.681, 0.685]** |
| Clinical DL | **0.790 [0.787, 0.793]** | 0.581 [0.578, 0.584] | **0.669 [0.667, 0.672]** | 0.683 [0.681, 0.685] | 0.682 [0.681, 0.685] |
| Text NB | **0.681 [0.678, 0.685]** | 0.647 [0.644, 0.650] | 0.664 [0.661, 0.666] | 0.719 [0.718, 0.721] | 0.711 [0.710, 0.714] |
| Text LR | 0.629 [0.626, 0.633] | 0.728 [0.724, 0.731] | 0.675 [0.672, 0.678] | 0.754 [0.752, 0.756] | 0.738 [0.736, 0.740] |
| Text RF | 0.514 [0.511, 0.517] | **0.783 [0.780, 0.787]** | 0.621 [0.618, 0.624] | 0.745 [0.743, 0.747] | 0.714 [0.712, 0.716] |
| Text GB | 0.578 [0.575, 0.581] | 0.758 [0.755, 0.762] | 0.656 [0.653, 0.659] | 0.753 [0.752, 0.755] | 0.732 [0.730, 0.734] |
| Text Net | 0.638 [0.635, 0.642] | 0.737 [0.734, 0.740] | **0.684 [0.681, 0.687]** | **0.760 [0.758, 0.762]** | **0.745 [0.744, 0.747]** |
| Global NB | **0.729 [0.726, 0.732]** | 0.635 [0.632, 0.638] | 0.679 [0.676, 0.681] | 0.720 [0.718, 0.722] | 0.715 [0.713, 0.717] |
| Global LR | 0.652 [0.649, 0.656] | 0.736 [0.733, 0.740] | 0.692 [0.689, 0.695] | 0.764 [0.762, 0.766] | 0.750 [0.748, 0.752] |
| Global RF | 0.585 [0.582, 0.589] | **0.776 [0.773, 0.779]** | 0.667 [0.665, 0.670] | 0.763 [0.761, 0.765] | 0.742 [0.740, 0.744] |
| Global GB | 0.616 [0.613, 0.620] | 0.762 [0.759, 0.765] | 0.681 [0.679, 0.684] | 0.766 [0.764, 0.768] | 0.748 [0.746, 0.750] |
| DeepEMC<sup>2</sup> | 0.671 [0.668, 0.675] | 0.742 [0.739, 0.745] | **0.705 [0.702, 0.707]** | **0.771 [0.770, 0.773]** | **0.759 [0.757, 0.761]** |
| $\Delta P$ (%) | 2.7 [2.1, 3.4] | 19.5 [18.8, 20.1] | 11.3 [10.7, 11.8] | 13.2 [12.9, 13.6] | 12.5 [12.1, 12.9] |
| $\Delta BM$ (%) | 1.9 [1.2, 2.6] | 0.6 [-0.1, 1.2] | 1.3 [0.7, 1.8] | 0.7 [0.4, 1.1] | 0.9 [0.5, 1.3] |

Abbreviations: NB, naive bayes; LR, logistic regression; RF, random forest; GB, gradient boosting; DL, deep learning; $\Delta P$ , DeepEMC<sup>2</sup> difference respect to the protocol; $\Delta BM$ , DeepEMC<sup>2</sup> difference respect to the best baseline model in life-threatening prediction (logistic regression).

**Table 4.** Performances of the in-house triage protocol, baseline models and deep learning models in response delay prediction (test set). Bootstrapped 95% confidence intervals are shown between brackets. Percentage differences between DeepEMC^2^—the global deep learning model—and the protocol ΔP (%), along with percentage differences between DeepEMC^2^ and the best baseline model ΔBM (%)—highest F1-score^MACRO^—are also reported.

| Model | Admissible response delay (undelayable, minutes, hours, days) |  |  |  |
| --- | --- | --- | --- | --- |
|  | Recall<sup>MACRO</sup> | Precision<sup>MACRO</sup> | F1-score<sup>MACRO</sup> | Accuracy |
| Protocol | 0.411 [0.409, 0.413] | 0.416 [0.414, 0.419] | 0.401 [0.398, 0.403] | 0.428 [0.426, 0.430] |
| Context NB | 0.375 [0.373, 0.377] | 0.382 [0.379, 0.385] | 0.364 [0.362, 0.366] | 0.396 [0.394, 0.399] |
| Context LR | 0.376 [0.374, 0.378] | 0.396 [0.393, 0.398] | 0.369 [0.367, 0.371] | 0.406 [0.403, 0.408] |
| Context RF | 0.348 [0.345, 0.350] | 0.357 [0.354, 0.359] | 0.350 [0.348, 0.352] | 0.371 [0.369, 0.373] |
| Context GB | **0.382 [0.380, 0.384]** | 0.414 [0.411, 0.417] | **0.383 [0.381, 0.385]** | **0.415 [0.413, 0.417]** |
| Context DL | 0.376 [0.374, 0.378] | **0.415 [0.412, 0.418]** | 0.377 [0.374, 0.379] | 0.413 [0.411, 0.415] |
| Clinical NB | 0.458 [0.456, 0.460] | 0.503 [0.501, 0.506] | 0.460 [0.458, 0.462] | 0.482 [0.480, 0.484] |
| Clinical LR | **0.479 [0.477, 0.481]** | 0.522 [0.520, 0.525] | **0.488 [0.486, 0.490]** | 0.505 [0.503, 0.507] |
| Clinical RF | 0.477 [0.475, 0.479] | **0.533 [0.530, 0.535]** | 0.485 [0.483, 0.488] | **0.507 [0.504, 0.509]** |
| Clinical GB | 0.477 [0.475, 0.479] | 0.532 [0.530, 0.535] | 0.485 [0.483, 0.488] | **0.507 [0.504, 0.509]** |
| Clinical DL | 0.477 [0.475, 0.479] | 0.530 [0.527, 0.532] | 0.485 [0.483, 0.487] | 0.506 [0.504, 0.508] |
| Text NB | 0.527 [0.524, 0.529] | 0.517 [0.515, 0.519] | 0.519 [0.517, 0.521] | 0.533 [0.531, 0.535] |
| Text LR | 0.544 [0.542, 0.546] | 0.564 [0.562, 0.567] | 0.550 [0.548, 0.553] | 0.569 [0.567, 0.572] |
| Text RF | 0.524 [0.522, 0.527] | **0.583 [0.581, 0.586]** | 0.535 [0.533, 0.538] | 0.563 [0.561, 0.566] |
| Text GB | **0.545 [0.543, 0.547]** | 0.577 [0.575, 0.580] | 0.554 [0.552, 0.556] | 0.574 [0.572, 0.576] |
| Text DL | 0.544 [0.542, 0.546] | 0.583 [0.580, 0.585] | **0.555 [0.553, 0.557]** | **0.576 [0.574, 0.578]** |
| Global NB | 0.537 [0.534, 0.539] | 0.531 [0.529, 0.534] | 0.533 [0.531, 0.535] | 0.549 [0.547, 0.551] |
| Global LR | 0.557 [0.555, 0.559] | 0.579 [0.577, 0.581] | 0.564 [0.562, 0.567] | 0.582 [0.580, 0.585] |
| Global RF | 0.547 [0.545, 0.549] | 0.593 [0.590, 0.595] | 0.557 [0.555, 0.560] | 0.581 [0.579, 0.583] |
| Global GB | 0.562 [0.560, 0.565] | **0.593 [0.591, 0.596]** | 0.572 [0.570, 0.574] | 0.589 [0.587, 0.592] |
| DeepEMC<sup>2</sup> | **0.569 [0.567, 0.571]** | 0.589 [0.587, 0.591] | **0.576 [0.574, 0.579]** | **0.592 [0.590, 0.594]** |
| $\Delta P$ (%) | 15.8 [15.4, 16.2] | 17.3 [16.8, 17.7] | 17.5 [17.1, 18.1] | 16.4 [16, 16.8] |
| $\Delta BM$ (%) | 0.7 [0.2, 1.1] | -0.4 [-0.9, 0] | 0.4 [0, 0.9] | 0.3 [-0.2, 0.7] |

Abbreviations: NB, naive bayes; LR, logistic regression; RF, random forest; GB, gradient boosting; DL, deep learning; $\Delta P$ , DeepEMC<sup>2</sup> difference respect to the protocol; $\Delta BM$ , DeepEMC<sup>2</sup> difference respect to the best baseline model in response delay prediction (gradient boosting).

**Table 5.** Performances of the in-house triage protocol, baseline models and deep learning models in jurisdiction prediction (test set). Bootstrapped 95% confidence intervals are shown between brackets. Percentage differences between DeepEMC^2^—the global deep learning model—and the protocol ΔP (%), along with percentage differences between DeepEMC^2^ and the best baseline model ΔBM (%)—highest F1-score^MACRO^—are also reported.

| Model | Emergency system jurisdiction (yes/no) |  |  |  |  |
| --- | --- | --- | --- | --- | --- |
|  | Single-class metrics (yes) |  |  | Two-class metrics (yes/no) |  |
|  | Recall | Precision | F1-score | Accuracy | F1-score<sup>MACRO</sup> |
| Protocol | 0.855 [0.854, 0.857] | 0.800 [0.798, 0.802] | 0.827 [0.825, 0.828] | 0.756 [0.754, 0.757] | 0.706 [0.703, 0.708] |
| Context NB | 0.892 [0.891, 0.894] | **0.752 [0.750, 0.754]** | 0.816 [0.815, 0.818] | 0.726 [0.724, 0.728] | **0.638 [0.636, 0.640]** |
| Context LR | 0.919 [0.918, 0.921] | 0.746 [0.744, 0.748] | 0.824 [0.822, 0.825] | 0.731 [0.729, 0.733] | 0.629 [0.627, 0.631] |
| Context RF | 0.850 [0.848, 0.852] | 0.745 [0.743, 0.747] | 0.794 [0.793, 0.796] | 0.699 [0.697, 0.701] | 0.618 [0.615, 0.620] |
| Context GB | 0.936 [0.935, 0.937] | 0.744 [0.742, 0.746] | 0.829 [0.828, 0.831] | **0.737 [0.735, 0.739]** | 0.628 [0.625, 0.630] |
| Context DL | **0.945 [0.943, 0.946]** | 0.741 [0.739, 0.743] | **0.830 [0.829, 0.832]** | 0.736 [0.734, 0.738] | 0.620 [0.618, 0.622] |
| Clinical NB | 0.897 [0.896, 0.899] | 0.800 [0.798, 0.802] | 0.846 [0.844, 0.847] | 0.777 [0.775, 0.778] | 0.720 [0.718, 0.723] |
| Clinical LR | 0.906 [0.904, 0.908] | 0.798 [0.796, 0.800] | 0.848 [0.847, 0.850] | 0.779 [0.777, 0.781] | 0.721 [0.718, 0.723] |
| Clinical RF | 0.901 [0.899, 0.902] | 0.801 [0.799, 0.803] | 0.848 [0.847, 0.850] | **0.780 [0.778, 0.782]** | **0.724 [0.722, 0.726]** |
| Clinical GB | **0.916 [0.914, 0.917]** | 0.793 [0.791, 0.795] | **0.850 [0.849, 0.851]** | 0.779 [0.778, 0.781] | 0.717 [0.714, 0.719] |
| Clinical DL | 0.900 [0.899, 0.902] | **0.802 [0.800, 0.804]** | 0.848 [0.847, 0.849] | **0.780 [0.778, 0.782]** | **0.724 [0.722, 0.726]** |
| Text NB | 0.793 [0.791, 0.795] | **0.833 [0.831, 0.835]** | 0.812 [0.811, 0.814] | 0.750 [0.748, 0.752] | 0.719 [0.717, 0.721] |
| Text LR | 0.896 [0.895, 0.898] | 0.810 [0.807, 0.811] | 0.851 [0.849, 0.852] | 0.785 [0.783, 0.787] | **0.734 [0.732, 0.736]** |
| Text RF | **0.936 [0.934, 0.937]** | 0.782 [0.780, 0.784] | 0.852 [0.851, 0.853] | 0.778 [0.776, 0.780] | 0.704 [0.702, 0.707] |
| Text GB | 0.906 [0.905, 0.907] | 0.803 [0.801, 0.805] | 0.851 [0.850, 0.853] | 0.784 [0.782, 0.786] | 0.728 [0.726, 0.730] |
| Text Net | 0.917 [0.916, 0.919] | 0.804 [0.802, 0.806] | **0.857 [0.856, 0.858]** | **0.791 [0.789, 0.793]** | **0.734 [0.732, 0.736]** |
| Global NB | 0.818 [0.817, 0.820] | **0.834 [0.832, 0.836]** | 0.826 [0.825, 0.828] | 0.765 [0.763, 0.766] | 0.731 [0.729, 0.733] |
| Global LR | 0.902 [0.901, 0.904] | 0.816 [0.814, 0.818] | 0.857 [0.855, 0.858] | 0.794 [0.792, 0.796] | 0.745 [0.743, 0.747] |
| Global RF | **0.925 [0.924, 0.926]** | 0.802 [0.800, 0.804] | 0.859 [0.858, 0.860] | 0.793 [0.791, 0.795] | 0.734 [0.732, 0.737] |
| Global GB | 0.914 [0.913, 0.916] | 0.811 [0.809, 0.813] | **0.860 [0.858, 0.861]** | 0.796 [0.794, 0.798] | 0.743 [0.741, 0.745] |
| DeepEMC<sup>2</sup> | 0.895 [0.894, 0.897] | 0.827 [0.825, 0.829] | **0.860 [0.858, 0.861]** | **0.801 [0.799, 0.802]** | **0.757 [0.755, 0.759]** |
| $\Delta P$ (%) | 4 [3.7, 4.3] | 2.7 [2.3, 3.1] | 3.3 [3, 3.6] | 4.5 [4.2, 4.8] | 5.1 [4.7, 5.6] |
| $\Delta BM$ (%) | -0.7 [-1, -0.4] | 1.1 [0.7, 1.5] | 0.3 [0, 0.6] | 0.7 [0.3, 1] | 1.2 [0.8, 1.6] |

Abbreviations: NB, naive bayes; LR, logistic regression; RF, random forest; GB, gradient boosting; DL, deep learning; $\Delta P$ , DeepEMC<sup>2</sup> difference respect to the protocol; $\Delta BM$ , DeepEMC<sup>2</sup> difference respect to the best baseline model in jurisdiction prediction (logistic regression).

### 4.1. Life-threatening level

Table 3 shows that DeepEMC^2^—the global DL model— highly outperforms the current protocol in the life-threatening prediction task with a 13.2% of accuracy improvement and a 12.5% of macro F1-score increment. This increment is statistically significant as reflected by the absence of overlapping in the 95% confidence intervals (CI). DeepEMC^2^ captures more true life-threatening situations—higher recall— being much more precise—with less false positives.

In comparison to the baseline models, although DeepEMC^2^ does not offer the best recall or precision, it achieves the best trade-off between them, as indicated by the best F1-score, being this metric statistically superior to those F1-scores attained by the baseline models. Likewise, referring to the best balanced two-class performance, DeepEMC^2^ presents the best macro F1-score, with statistically significant difference respect to the baselines models.

Focusing on the subnetworks, the ConNet is the weakest deep learning model. The CliNet offers the better detection rate for true life-threatening situations but at the expense of a significant amount of false positives. Finally, the TextNet exhibits the overall better behavior although its capability to capture true life-threatening events is not the best among the subnetworks.

Regarding the comparative performance among the subnetworks and their respective baseline models, it stands out the performance similitude among the ConNet and some of their associated baseline models as well as the high outcomes resemblance among the CliNet and the baseline models using clinical variables. Finally, notably the TextNet presents greater differences respect its corresponding baseline models, being these differences notorious in the F1-score and macro F1-score.

### 4.2. Admissible response delay

Table 4 shows that DeepEMC^2^ outcomes are significantly superior to those achieved by the protocol in the response delay prediction task (CI 95%).

Overall detection of situations with a specific admissible response delay (undelayable, minutes, hours, days) is largely improved by DeepEMC^2^—15.8% increment in macro recall—while remarkably enhancing overall precision —17.3% increment. Regarding the general performance in all classes, DeepEMC^2^ significantly improves the protocol, with a 16.4% of accuracy improvement and a 17.5% of macro F1-score increment.

DeepEMC^2^ does not offer the best overall precision compared to the baseline models. However, it improves the overall recall and the best balanced multiclass performance, in terms of macro F1-score. Furthermore, this global performance is the best, in terms of statistically significance difference respect the baseline models, although the performance difference respect the global gradient boosting model—best baseline model in admissible response delay prediction—is at the limit, since 0 is the lower bound of the 95% confidence intervals for performance differences.

Focusing on DeepEMC^2^ subnetworks for response delay prediction, the ConNet is at the bottom in performance terms, not being capable of outperforming the protocol. The CliNet is clearly over the ConNet and already beats the protocol, while the TextNet is the best DeepEMC^2^ subnetwork in all metrics, with a substantial increase respect to the CliNet.

Regarding the comparative performance among the subnetworks and their respective baseline models, it can be appreciated the performance similitude among the ConNet and some of their associated baseline models as well as the high outcomes resemblance among the CliNet and the baseline models fed with the clinical variables. Finally, the TextNet presents greater differences respect its corresponding baseline models, being these differences significant in the macro F1-score metric.

### 4.3. Emergency system jurisdiction

Table 5 shows that DeepEMC^2^ significantly outperforms the protocol in the jurisdiction prediction task (95% CI). It captures more situations which are jurisdiction of the emergency system—better recall—being more precise—with less false positives. Respect to the overall performance in both classes, DeepEMC^2^ surpasses the protocol, with a 4.5% of accuracy improvement and a 5.1% of macro F1-score increment.

DeepEMC^2^ does not offer the best recall or precision compared to the baseline models. However, it achieves, along with the gradient boosting model, the best trade-off between them, as indicated by their best F1-score, being this metric statistically superior to that attained by the logistic regression model—best baseline model in emergency system jurisdiction prediction. Likewise, referring to the best balanced two-class performance, DeepEMC^2^ presents the best macro F1-score, with statistically significant differences respect the baselines models.

Focusing on DeepEMC^2^ subnetworks, although the ConNet presents the highest recall values, its precision is not the best, with worse general results than the protocol in the jurisdiction prediction task. The CliNet provides a substantial improvement over the later subnetwork, with an overall performance above the protocol. As in life-threatening and response delay, the TextNet is the subnetwork attaining the best outcomes.

Regarding to the comparative performance among the subnetworks and their respective baseline models, notably the performance is similar among the ConNet and some of their associated baseline models as well as the high outcomes resemblance among the CliNet and the baseline models fed with the clinical variables. Finally, it has to be highlighted that the TextNet presents greater differences respect its corresponding baseline models, being these differences notorious in the F1-score and accuracy.

## 5. Discussion

### 5.1. Relevance

The superior performance of DeepEMC^2^ and some of the baseline models, respect to the in-house triage protocol, suggests the existence of information provided during the emergency medical call not considered by the current protocol, but captured by the machine learning models. Likewise, the DL approach is preferable over the other families of models tested, since DeepEMC^2^ outcomes are significantly above those attained by the baseline models.

In referring context and clinical variables, DL is not clearly at the top. However, regarding the free text dispatcher observations, the DL approach is, overall, remarkably superior. Likewise, as TextNet outcomes are far better than those attained by the ConNet and CliNet, the most valuable information provided during the emergency medical call would be present at these unstructured features. Since text fields are unbounded, they would embrace wider casuistry, allowing more precision in the EMCI description, lowering, consequently, its uncertainty.

Regarding the clinical variables, they stand as an excellent life-threatening detector features—about 80% of total cases. This could be due to the fact that dispatchers ask for them to reduce chances of missing situations where patient’s life is at risk. Similarly, the outstanding emergency system jurisdiction recall of demographics and circumstantial factors—capturing about 95% of total cases—may be related with patient profiles highly susceptible from requiring emergency aid, e.g., elderly cardiac patient males.

Comparing classification scores across tasks, the hardest classification problem appeared to predict the admissible response delay, probably derived from the fact that it is a multiclass label, presenting twice possible outputs (undelayable, minutes, hours, days) than the other labels (life-threatening, jurisdiction), which are binary.

The modular approach followed in this work, assembling four specialized subnetworks into a single global network (DeepEMC^2^), has shown that the potential of the aggregated network is superior to any of its individual components, balancing their respective weaknesses and strengths while properly integrating processed information within each one.

Finally, the results of this work imply that current emergency dispatch processes could be improved by means of deep learning, eventually deriving in a positive impact over patient wellbeing and health services sustainability.

### 5.2. Limitations

The main limitation of this work is the inherent uncertainty bound to the problem: in the studied dataset it was likely to find similar input combinations presenting completely different label values. In other words, the challenge faced in this work exhibits classes overlap, where different disorders may present the same clinical picture. For example, chest pain may imply a life-threatening situation, if the underlying unknown cause is a heart attack, or not, since it could be derived from a prior anxiety crisis. This non-discriminative variability sets bounds in terms of maximum performance attainable by any model—Bayes error [95].

Besides, the data available to conduct this work lies between 2009 and 2012 years (both included). Even though the clinical framework of pathologies like heart failure or epileptic crisis could be fairly constant across time, an in-depth study of potential dataset shifts [96,97], and related abrupt or gradual changes regarding the statistical distributions of new data [98] has to be carried out before implementing the model in emergency medical dispatch centers.

### 5.3. Future work

Next steps include the evaluation of DeepEMC^2^ with prospective cases from the Valencia region—with more recent incidents, monitoring the aforementioned dataset shifts and acting in consequence. Passing this phase favorably would enable us to begin the integration of the model in an emergency medical dispatch center, with a prospective evaluation of the system performance and added value on routine settings through a randomized controlled trial for CDSS [99,100]. To accomplish that, a graphical user interface will be proposed, to allow the interaction between the dispatcher and the model during the call. Finally, the resulting tool will be implemented in the emergency medical dispatch center of the Valencian Community.

## 6. Conclusions

A novel deep ensemble multitask model (DeepEMC^2^) designed to aid non-clinical dispatchers during emergency medical calls to classify incidents by their life-threatening level, admissible response delay and emergency system jurisdiction, has been developed and successfully evaluated. To our knowledge, this is the first deep learning model implemented to face this challenge.

The performance achieved by the model is highly superior to that attained by the current in-house triage protocol of the emergency medical dispatch service of the Valencian Community, achieving a macro F1-score improvement of 12.5%, 17.5%, 5.1% in life-threatening, response delay and jurisdiction classification, respectively. Likewise, DeepEMC^2^ outcomes are above those accomplished by the additional machine learning models tested, including naive bayes, logistic regression, random forest and gradient boosting. This increment was proved as statistically significant (α=0.05).

Remarkably, the network modular design with specialized subnetworks for the different data modalities has allowed discovering the potential benefit of the information contained in free text fields for the automatic classification of emergency medical call incidents. This information can be used to optimize current guidelines.

The implantation of this model in medical dispatch centers would have a remarkable impact in patient wellbeing and health services sustainability.

## Data Availability

The data use is restricted by the Valencian Agency for Security and Emergency Response.

## Funding

This work has been supported by the *Valencian agency for security and emergency response* project A180017304-1, the *Ministry of Science, Innovation and Universities* of Spain program FPU18/06441 and the *EU Horizon 2020* project InAdvance 825750.

## Declaration of competing interest

The authors declare no competing interests.

## Acknowledgements

We thank the support of physicians from the Health Services Department of Valencian Community. CS acknowledges the support of NVIDIA GPU Grant Program.

## Notes

### Competing Interest Statement

The authors have declared no competing interest.

### Funding Statement

This work has been supported by the Agència Valenciana de Seguretat i Resposta a les Emergències project A180017304-1, the Ministerio de Ciencia, Innovación y Universidades of Spain program FPU18/06441 and the EU Horizon 2020 project InAdvance 825750.

### Author Declarations

Participation consent protocols, as provided by the Health Services Department of the Valencian Community.

### Summary of Updates

Inclusion of the following baseline models: naive bayes, logistic regression, random forest and gradient boosting. This allows a more exhaustive evaluation of the results. In addition, materiales from the supplementary data file have been moved to the main manuscript. New metrics, such as the macro F1 score have been included.

